# Resolving the Genomic Context of Clinically Relevant Antibiotic Resistance Genes in Wastewater with Ligation-Mediated PCR

**DOI:** 10.64898/2026.09.09.26361541

**Authors:** Megan E. O’Brien, Bradie Ahern, Piper Brase, Sooyeol Kim, Caroline E.M. McCormack, Denise Garcia, Erika Keim, Erin Dahl, Matthew Feck, Kelly Kauber, Jorge A. Marchand, Rose S. Kantor, Amy J. Pickering, Breanna McArdle, Erica R. Fuhrmeister

**Affiliations:** Department of Environmental and Occupational Health Sciences, University of Washington, Seattle, Washington, USA; Department of Civil and Environmental Engineering, University of Washington, Seattle, Washington, USA; Washington State Department of Health, Office of Communicable Disease Epidemiology, Shoreline, Washington, USA; Department of Civil and Environmental Engineering, University of California, Berkeley, Berkeley California, USA; School of Civil Engineering, the University of Sydney, Sydney, NSW, Australia; Washington State Department of Health, Environmental Laboratory Servies, Shoreline, Washington, USA; Department of Chemical Engineering, University of Washington, Seattle, Washington, USA; Physical and Life Sciences Directorate, Lawerence Livermore National Lab, California, USA; Biohub, San Francisco, CA, USA

## Abstract

Antimicrobial resistance (AMR) is a pressing global public health challenge. AMR is driven in part by the spread of antibiotic resistance genes (ARGs) through bacterial communities via mobile genetic elements. Influent wastewater is a promising sample type for monitoring AMR because it pools biological inputs shed by individuals across a population. However, untargeted sequencing approaches, such as metagenomic sequencing, often miss low-abundance targets such as clinically important ARGs. In this work, we develop a ligation-mediated PCR (LM-PCR) enrichment strategy that can directionally capture the genomic context surrounding an ARG using long-read sequencing. We applied this method to study the natural genomic context diversity of four clinically relevant ARGs (*bla*_CTX-M_, *bla*_KPC_, and *bla*_OXA-48-like_, and *qnrS*) across 13 wastewater treatment plants in Washington state, each sampled at two timepoints. Across all timepoints, LM-PCR identified distinct genomic context cluster families associated with each ARG, including seven for *bla*_KPC_, 11 for *bla*_CTX-M_, 24 for *qnrS*, and one for *bla*_OXA-48-like_. Notably, 11 of the 24 *qnrS* containing clusters were putatively novel, with no matches to existing sequences in public databases. Genomic contexts associated with *bla*_CTX-M_ and *bla*_KPC_ were comparatively conserved across clusters, whereas *qnrS* was associated with a more diverse set of genetic sequences. Together, these results demonstrate that LM-PCR can resolve low-abundance, ARG-associated genomic variation in complex wastewater samples and provide a scalable framework for tracking the dissemination of clinically relevant AMR determinants.

## 1.0 Introduction

Antimicrobial resistance (AMR) is one of the most pressing global health threats recognized by the World Health Organization as a top priority for public health action.^1^ By 2050, a projected 1.9 million deaths per year are expected to be directly caused by AMR infections, with an additional 8.2 million deaths expected to be caused by closely-associated illnesses.^2^ The Centers for Disease Control and Prevention (CDC) estimates the United States spends $4.6 billion,^3,4^ heavily rely on clinical data provided by human and veterinary medicine. By design, this monitoring strategy is reactive rather than proactive, as it relies on data generated after resistant infections have already occurred. Although essential for patient treatment, this approach provides limited insight into the population-level dynamics and transmission of antibiotic resistance genes (ARGs).

Wastewater-based epidemiology (WBE) is an environmental monitoring strategy that has historically been used for surveillance of polioviruses,^5^ enteroviruses,^6^ SARS-CoV-2,^7–9^ and various other pathogens that can be excreted in urine, mucus, and feces.^10^ The COVID-19 pandemic launched a substantial interest in WBE, with public health agencies, schools, and even healthcare facilities publicly disseminating findings in wastewater through research and data dashboards.^11–15^ The same population-level sampling that makes WBE valuable for pathogen surveillance also makes it well suited for monitoring the emergence and spread of AMR. Specifically, ARGs can be present in an individual without an active resistant infection, and that resistance can be transferred to other humans or animals,^16–18^ making population-level surveillance useful for capturing resistance that clinical, infection-based monitoring misses.

Previous investigations of ARGs in influent wastewater have mostly relied on amplicon-based detection.^19–21^ Although amplicon-based approaches can detect ARGs of interest with high sensitivity, they often lose the surrounding genomic context needed to determine how these genes are mobilized, transmitted, and maintained across microbial populations. The genomic context of ARGs can contain critical information regarding mobility and host association, including the presence of mobile genetic elements (MGEs), prophage-associated regions, and neighboring sequence features that help identify the putative microbial host range of an ARG. These features can reveal whether ARGs are linked to elements capable of horizontal gene transfer (HGT) among pathogenic and non-pathogenic microorganisms, information that ARG prevalence alone cannot provide.^22,23^ Characterizing genomic context surrounding ARGs can help us better understand how ARGs are mobilized and disseminated across bacterial populations.^24^

Capturing the genomic context of ARGs is particularly important in wastewater, which is a known hotspot for AMR transfer and dissemination.^22,25,26^ Metagenomic sequencing of ARGs in influent wastewater has grown in recent years and has contributed to a greater understanding of the global resistome.^27–29^ However, short-read sequencing strategies often struggle to assemble ARG-containing regions since identical or highly similar ARG sequences can occur in multiple genomic contexts.^30,31^ Alternatively, long-read sequencing strategies can overcome these limitations as they directly sequence long fragments of DNA that can stretch across repetitive regions.

For both long- and short-read metagenomic sequencing approaches, clinically important ARGs are often too low in abundance to obtain adequate sequencing depth.^32,33^ Targeted sequencing strategies have recently emerged to address these limitations. Methods including probe-based capture^34,35^ are highly accurate and compatible with a wide range of sample types and targets, but they remain resource intensive, making them unsuitable for routine monitoring. A lower-cost, resource-efficient sequencing strategy that enriches clinically important ARGs while preserving their genomic context would make it possible to expand surveillance across more communities, environments, and sampling timepoints.

To meet this need, we developed a modified ligation-mediated PCR (LM-PCR) enrichment approach that selectively amplifies ARG-containing DNA fragments and is compatible with current long-read sequencing technologies. This method enables effective recovery of extended genomic regions surrounding low-abundance ARGs. We characterized the genomic contexts of four clinically relevant ARGs (*bla*_CTX-M_, *bla*_KPC_, *bla*_OXA-48-like_, and *qnrS)* in 13 wastewater treatment plants (WWTPs) across Washington (WA) state at two timepoints one month apart. To evaluate performance of influents with variable characteristics, we selected WWTPs with differing service populations and land-use characteristics. Additionally, we also evaluated diversity in genomic contexts by gene and whether common sequence patterns were observed across multiple site populations.

The ARGs targeted in this study are of clinical significance due to their association with resistance to broad-spectrum and last-resort antibiotics, as well as their potential for horizontal transfer. *Bla*_CTX-M_, *bla*_KPC_, and *bla*_OXA-48-like_ confer resistance to β-lactam antibiotics,^36–38^ including third-generation cephalosporins and carbapenems, while *qnrS* mediates resistance to fluoroquinolones.^39^ These genes are commonly located on MGEs that facilitate transmission among bacterial populations across human, animal, and environmental reservoirs.^40–42^ Monitoring their occurrence in wastewater influent can provide insight into the community-level prevalence of clinically important ARGs. Characterizing their genomic context can further elucidate the mechanisms underlying their dissemination.

## 2.0 Methods

### 2.1. Culturing Positive Control Bacterial Isolates

Bacterial species harboring ARGs of interests were obtained from the CDC Antimicrobial Resistance Isolate Bank (**Table S1**) and served as positive controls for method development. Isolates were grown by inoculating 15 mL of LB broth with 15 μL of glycerol stock and incubated at 1,300 RPM overnight at 37°C. The culture was pelleted at 5000 x g and genomic extractions were conducted using the NEB Monarch Spin Genomic DNA Purification Kit (Cat. T3010S) and DNA was quantified using DeNovix Fluorometer with Qubit 1X dsDNA High Sensitivity assay kit (Invitrogen). Isolate extracts were confirmed to have selected targets through qPCR (**Table S2**). Genomes were obtained for each isolate and used as references for read mapping.

### 2.2 Assay and Primer Design

To develop our enrichment method, a DNA mixture with equal amounts of three bacterial isolates from CDC’s Antibiotic Resistance (AR) Isolate Bank were prepared containing the targets *bla*_KPC_, *bla*_CTX-M_, *bla*_OXA-48-like_, and *qnrS* (**Table S1**). Our enrichment strategy was based on a modified ligation-mediated PCR (LM-PCR) workflow. Briefly, a Y-shaped adapter was ligated to the ends of DNA fragments in each sample. During PCR, the ARG-specific forward primer amplifies the ARG and the proceeding genomic context. This process creates the reverse compliment of the Y-adapter sequence at the end of the fragment, to which the Y adapter primer will bind. Thus, to achieve exponential amplification, the strand must contain both the ARG and the Y adapter (**Figure 1**).

**Figure 1:**
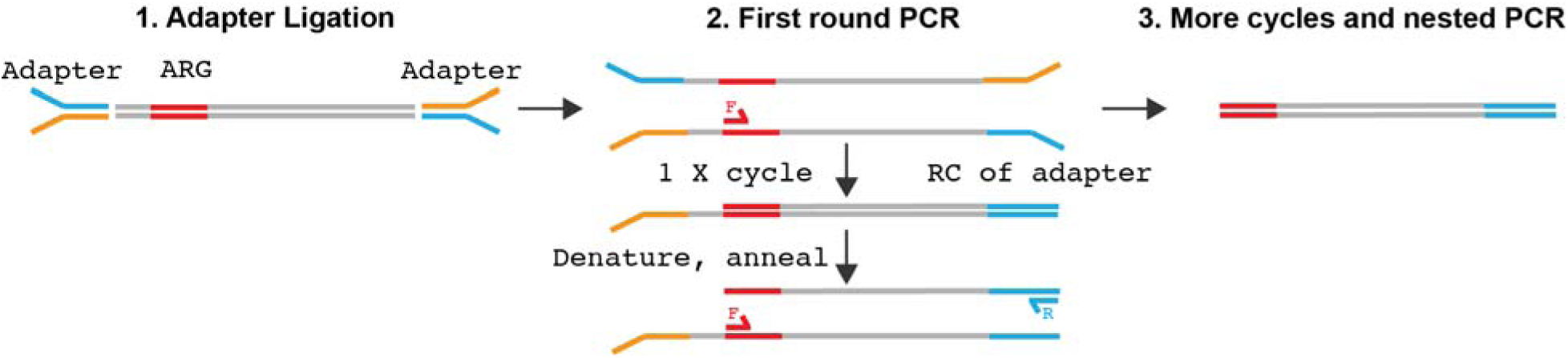
Schematic for amplification of ARG and genomic context. A known DNA adapter sequence is ligated to the end of all DNA fragments in a sample (Step 1). A PCR-reaction with two primers (1 ARG primer and 1 adapter primer) amplifies all fragments containing the ARG and the adapter (Step 2). This ensures that we amplify only the targets of interest, as exponential amplification can only occur if a sequence has both an ARG and an adapter sequence (Step 3).

Forward primers were designed to target the 5’ region of the ARG (within the first 150 bps) using the PrimerQuest tool (Integrated DNA Technologies). The primer region was chosen to capture as many alleles in the CARD database^44^ as possible with a single primer (**Table S2**). We performed multiple validation experiments including sequencing positive controls, comparing multiplex to singleplex amplification, and testing the method on five unique wastewater samples.

### 2.3 Site Selection

We selected wastewater influent samples to represent a diverse range of populations and locations across WA state. All land-use data were self-reported by WWTPs and collected by the Washington State Department of Health’s Wastewater-Based Epidemiology Program (WAWBE). Vaccination data were obtained from WA State School Immunization Dashboard^43^ for the 2024-2025 school year. Data is reported at the school level as the percentage of enrolled K– 12 students with complete immunizations (MMR, Tdap, varicella, hepatitis B). Because each sewershed serves multiple schools, coverage is reported as the range of school-level percentages across schools within the catchment area of the wastewater treatment plant.

### 2.4 Sample Collection

24-hour composite influent samples were collected from 13 WWTPs (**Table 1)** in Washington state during the month of May and June 2025 (26 total samples). Each WWTP shipped 250 mL of wastewater in sterile HDPE bottles at 4°C to the Washington State Public Health Laboratories. Samples were picked up the next day and transported to the University of Washington, Seattle, and stored at 4°C overnight if processing was not conducted on the same day as pickup.

**Table 1:** Wastewater treatment plants (WWTPs) sampled, influent and sewershed characteristics. Population served by the WWTP, WWTP flow capacity (MGD), sewershed density in people per acre, self-reported survey responses for agricultural inputs, vaccination percentages in schools, number of hospitals in the catchment area, WA State Emergency Preparedness Region.

| Site | Population | Flow Rate | Sewershed Density per acre | Peri-Domestic Animals | Dairy Milk Processing | Meat Processing | Vaccination % Range<sup>2</sup> | # Hospitals | Emergency Preparedness region |
| --- | --- | --- | --- | --- | --- | --- | --- | --- | --- |
| WWTP 1 | >100,000 | >20 | 12.57 | Yes | No | No | 49-100 | 13 | 6 |
| WWTP 2 | >100,000 | >20 | 4.86 | Yes | Yes | Yes | 60-96 | 7 | 9 |
| WWTP 3 | >100,000 | >20 | 6.70 | NR<sup>1</sup> | NR | NR | 87-93 | 2 | 1 |
| WWTP 4 | >100,000 | 5-10 | 4.36 | Yes | No | Yes | 59-92 | 1 | 9 |
| WWTP 5 | >100,000 | 5-10 | 5.30 | No | No | No | 80-98 | 1 | 8 |
| WWTP 6 | 20,001-100,000 | 1-5 | 5.72 | No | No | Yes | 67-88 | 1 | 1 |
| WWTP 7 | 20,001-100,000 | 5-10 | 4.41 | Yes | No | Yes | 73-97 | 2 | 8 |
| WWTP 8 | 20,001-100,000 | 1-5 | 3.99 | NR | NR | NR | 60-87 | 0 | 1 |
| WWTP 9 | 20,001-100,000 | 1-5 | 3.41 | Yes | No | No | 80-92 | 1 | 7 |
| WWTP 10 | 10,000-20,000 | 1-5 | 3.40 | NR | NR | NR | 88-97 | 0 | 8 |
| WWTP 11 | 10,000-20,000 | 1-5 | 2.34 | No | No | No | 61-78 | 1 | 2 |
| WWTP 12 | <5000 | <1 | 1.31 | Yes | No | No | 90-100 | 1 | 7 |
| WWTP 13 | <5000 | <1 | 2.58 | No | No | No | 77-84 | 1 | 1 |

<sup>1</sup>: NR: Not reported.
<sup>2</sup>: % of K-12 students fully vaccinated for all required vaccines (MMR, Tdap, Varicella, Hep B)

### 2.5 Wastewater Concentration and DNA Extraction

50 mL of influent was centrifuged at 16,000 x g for 12 minutes in a 50 mL conical tube. The supernatant was immediately decanted, leaving the pellet in the conical. This process was repeated until a total of 200 mL was concentrated. The pellet was resuspended in 1 mL of PBS and mixed thoroughly by vortexing. DNA was extracted from 250 µL of the resuspended pellet with the Qiagen AllPrep PowerViral Kit (Cat. 28000-50) according to manufacturer’s instructions and eluted in 50 µL of nuclease free water. Extracts were stored at −20°C until enrichment. A nucleic acid extraction blank was included with each batch. DNA concentrations were quantified using a Qubit and quality checked using a nanodrop spectrophotometer.

### 2.6 PCR Enrichment and qPCR Validation

200 ng of extracted DNA was dA-tailed with 3 μL of End Repair enzyme (NEB cat. #E7646) and 7 μL End Repair Buffer (NEB cat. # E7647) to a total volume of 50 μL in nuclease free water and incubated at 25°C for 5 minutes. The enzymes whereas then inactivated by incubating at 65°C for 5 minutes. The end-prepped sample was then cleaned using magnetic beads (Sergi Lab Supplies, cat. #1040) using a ratio of 0.4X (20 μL beads/50μL sample), which selects for fragments larger than 1000 bp. Using a magnetic bead rack, the resulting supernatant was removed and the beads were rinsed twice with freshly prepared 80% ethanol and eluted in 41 μL in nuclease free water. DNA was then quantified with a Qubit and diluted to 50 ng in 40 μL of nuclease free water.

Concurrently, the Y-adapter mix was prepared with 175 µL of nuclease-free water, 25 µL of each 100 μM Y-adapter oligo, and 25 µL of 10X T4 DNA ligase buffer (NEB cat. # B0202S) and heated at 95°C for 5 minutes. To ensure proper adapter annealing, we waited at least 15 minutes for the Y-adapter mix to cool at room temperature (approximately 21°C) before continuing to the ligation step. Then 10 µL of the cooled Y-adapter mix was added to the 40 μL sample and pipette mixed with 50 μL of 2X blunt/TA Mastermix (NEB, cat. # M0367S). The reaction was incubated for 15 minutes at 25°C. The cleanup procedure is as previously described (0.4X bead ratio with ethanol washes) and eluted in 50 µL nuclease free water.

Nested PCR was used to reduce non-specific amplification of our quadraplexed targets. A total of four ARG primers (for *bla*_KPC_, *bla*_CTX-M_, *bla*_OXA-48-like_, and *qnrS*) were combined in the PCR reaction with an equimolar amount of the Y-adapter primer to each ARG primer (**Table S3**).

In brief, 10 ng of adapter-ligated DNA was enriched using the first set of quadraplexed primers and amplified for 20 cycles. Then DNA was size selected (>1000 bp) as described above. A second round of PCR was conducted using a different set of quadraplexed ARG primers with barcodes for an additional 25 cycles (**Tables S4** and **S5**). The same DNA cleanup protocol as described above was used prior to qPCR and sequencing.

Before sequencing, all samples were quantified before and after PCR enrichment with qPCR. We determined 10^10^ gc/µL in the enriched sample was sufficient for sequencing. Additional details are available in the supporting information **Tables S6-S8**.

### 2.7 Nanopore Sequencing

PCR primers used during the second round of PCR contained Oxford Nanopore Technologies (ONT) nanopore sequencing barcodes (**Table S3**) for sample multiplexing. Approximately 200 ng of each sample was pooled and 4-6 samples were run on one MinION R10.4.1 flow cell. Libraries were prepared using the Ligation Sequencing Kit (SQK-LSK V14) according to manufacturer’s instructions with modifications. The DNA repair step was eliminated to reduce formation of concatemer products. Samples were loaded onto MinION R10.4.1 Flow Cells and were run for 72 hours. Sequencing data is available on the NCBI Sequence Read Archive (SRA) database under BioProject PRJNA1514801 (**Table S9**).

### 2.8 Bioinformatics Analyses

#### 2.8.0 Overview

Our bioinformatic pipeline used a conservative strategy to cluster and polish reads obtained from multiplexed nanopore sequencing. In this pipeline, raw nanopore reads were basecalled and processed to recover demultiplexed, sample-specific clusters of ARG-flanking sequences. Basecalled reads were length-filtered, split (internal barcode, adapter, or primer junctions) to resolve concatemeric and chimeric products, and demultiplexed by sample barcode. Resulting demultiplexed reads were retained only if they contained one of the expected ARG targets at either end of the sequence. For each sample and ARG, the filtered reads were then clustered to generate polished consensus sequences. Consensus sequences from each sample were dereplicated across the full dataset to retain one representative consensus sequence per cluster variant. The resulting representative consensus sequences were compared to sequences found in public databases for annotation. Sequences identified in multiple sample locations or timepoints, but without nearly identical hits in public databases, were labeled ‘novel’ and annotated using the nearest matching sequences. A more detailed description of the bioinformatic pipeline is provided below. The full bioinformatic pipeline is available on github (fuhr-microlab/lmpcr_arg).

#### 2.8.1 Basecalling and barcode Analysis

Raw nanopore signals were first basecalled using the Oxford Nanopore Technology (ONT) Guppy basecaller (v6.5.7, dna_r9.4.1_450bps_fast) with a read quality threshold of Q7. Resulting sequences were then length filtered to retain fragments (>1kbp for wastewater samples, >1.5kbp for positive controls) using seqtk (v1.2-r94). Prior to demultiplexing, reads were screened for internal (>200 bp from ends of reads) barcode or Y-adapter sequences using BLASTn (NCBI, BLASTn-short task; word size 4; identity ≥80%; query coverage ≥90%; E-value ≤10^-5^, gap-open penalty 5; gap-extend 2; mismatch penalty −1; match reward +1; minimum query overlap 80%). Reads containing internal matches were split and resulting read fragments shorter than 1,000 bp were discarded. Reads were then demultiplexed using cutadapt (v5.0) based on the one-sided barcode unique to each sample site (**Table S3**) with the following parameters: error rate 0.2; minimum adapter overlap 14 nt; no trim. A subsequent BLASTn search with a minimum query overlap 90% was used to remove concatemers. Remaining reads were mapped to ARG reference sequences to confirm that each read started or ended within the expected forward primer window by BLASTn (identity ≥60%; minimum ARG overlap 50%).

#### 2.8.2 Read clustering and polishing

For each sample barcode, filtered reads were grouped by ARG and clustered using VSEARCH (v2.21.1, cluster_fast; sequence identity threshold 0.85; query coverage 0.9; target coverage 0.9; both strands evaluated). Reads within each retained cluster were aligned to the longest cluster member as a draft reference using minimap2 (map-ont preset; no secondary alignments). Clusters containing fewer than 15 reads were discarded to minimize low-quality polishing. Polished cluster consensus sequences were generated using RACON (v1.5.0; match +3; mismatch −5; gap −4; window length 500 bp; quality threshold 10; up to 50 reads per cluster for large clusters).

#### 2.8.3 Unique gene cluster identification and gene annotation

RACON-polished clusters were dereplicated across the entire dataset by mapping each polished cluster to an external database using megaBLAST (word size 28; E-value ≤10^-10^). Here, the full NCBI nt database was used rather than the core nucleotide (core_nt) database in order to capture ARG-bearing sequences held in plasmid, environmental, and metagenomic records that are absent from core_nt. Polished clusters that best mapped the same external species (as determined by bitscore) with high agreement (>95% ID, >95% query coverage) were assigned to the same cluster group and numbered. For each unique numerical cluster within each of the ARG sets, the longest polished consensus sequence was chosen as the representative ARG gene cluster for downstream annotation. Representative cluster sequences identified using this strategy closely matched the sequences observed the external NCBI nt database (%ID: median = 99.84%; max 100%; min = 96.35%. % query coverage: median = 99.14%, max = 100%; min = 95.22%). Gene annotations from the best external database hit (as determined by bitscore) were used for annotating these representative clusters.

A subset of RACON-polished cluster sequences did not map to entries in the NCBI nt database at the previously set conservative threshold (>95% ID, >95% query coverage). An all-by-all BLASTn between these sequences was performed. Sequences that matched (>95% query coverage, >95% identity) in at least two samples (either two locations or two time points) were labeled as candidate ‘novel’ gene clusters and numbered. For these ‘novel’ gene cluster candidates, the longest polished consensus sequence was chosen as the representative sequence for downstream annotation. To annotate the resulting representative ‘novel’ gene cluster sequences, the ARG sequences were separated from the downstream sequence context and used in separate megaBLAST queries on an external database (NCBI nt). Annotations from the best hit (as determined by bitscore) were used for annotating the genes on these sequences.

To determine if our representative cluster sequences were present in isolates previously observed in Washington state, we mapped our sequences (BLASTn; 90% ID, 90% query coverage) against publicly available genomes from NCBI’s Pathogen Detection Project (Accessed July 2026).^44^ For this analysis, only genomes from *Salmonella enterica*, *E. coli* and *Shigella* spp., and *Klebsiella pneumoniae* that included Washington state (WA, USA) in their location metadata were used.

### 2.9 Data Analysis

To compare abundance between timepoints we calculated the Pearson correlation coefficient. For ordination, we calculated Bray-Curtis dissimilarity and conducted a PCoA using cmdscale in R v4.3.2. Clustering of samples in the PCoA was determined using partitioning around medoids and average silhouette width with the cluster library (v2.1.6). We tested the contribution of population served, flow rate, sewershed density, vaccination range in public schools, number of hospitals, and emergency preparedness region to the variance using PERMANOVA with adonis function in R using the vegan library (v2.6.4).

## 3.0 Results

### 3.1 Protocol Optimization

We first tested our multiplex enrichment method on three pooled *Klebsiella pneumoniae* isolates carrying all four ARG targets: *bla*_CTX-M_, *bla*_OXA-48-like_, *bla*_KPC_, *qnrS*. Target-specific read recovery was highest for *bla*_KPC_ (62.3% of reads >1000 bp) and *bla*_CTX-M_, (38.1%), with lower recovery for *bla*_OXA-48like_ (3.3%) and *qnrS* (2.6%) (**Table S11**). The reduced recovery of *qnrS* for the positive control likely reflects target-specific amplification bias. In the control, *qnrS* and *bla*_CTX-M_ genes are separated by only 4,616 bp in the reference isolate, and *bla*_CTX-M_ was preferentially amplified under these multiplex conditions. The annotations for the genomic context surrounding these positive control samples match the reference *K. pneumoniae* isolate sequences (**Table S1**, **Figure 2**). This positive control experiment also facilitated pipeline development of the bioinformatic pipeline to demultiplex, cluster, and polish reads.

**Figure 2:**
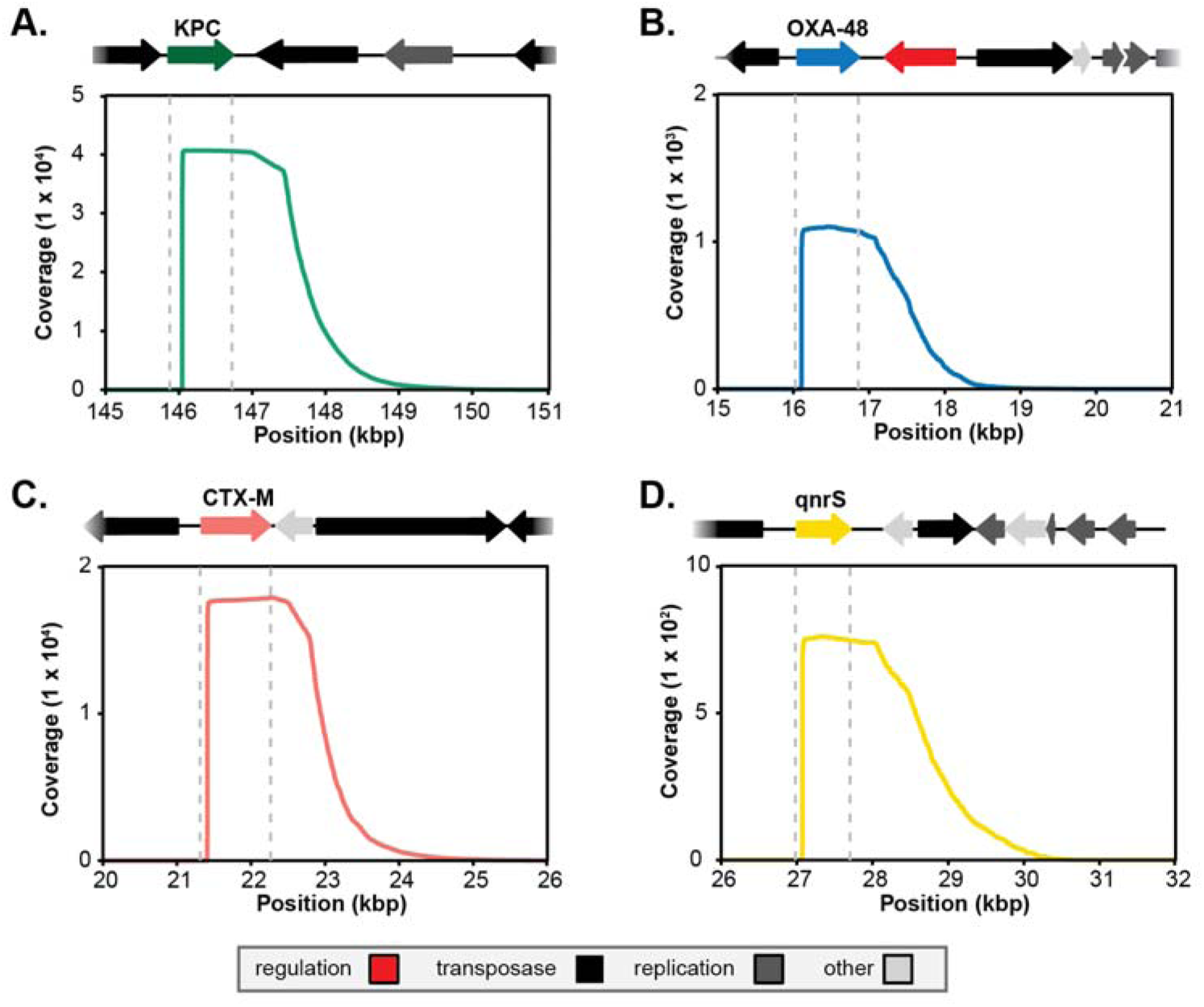
Multiplexed ligation-mediated PCR enrichment of ARGs and downstream genomic context from Klebsiella pneumoniae isolates. Genomic annotation and coverage of positive controls for **(A)** bla_KPC_ **(B)** bla_OXA-48-like_, **(C)** bla_CTX-M_, and **(D)** qnrS. All annotated sequences matched to the corresponding control isolates (Table S1). Samples were not digested prior to sequencing; all fragmentation is natural shearing. Coverage ranged from approximately 7×10^2^ **(D)** to 4×10^4^ **(A)**, depending on the target. The vertical dotted gray lines represent the length of the specific gene in the isolate genome. The colored line depicts read coverage.

To ensure our ARG enrichment method worked at environmentally relevant concentrations, we then tested our method on all four targets across five different wastewater samples. For all samples, the ARGs of interest alongside their downstream contexts were successfully enriched using a quadplex LM-PCR assay (**Figure S1**). Of the final processed reads >1,000 bp in length, the majority aligned to our four targets (**Table S12**). No target reads were observed in our untargeted sequencing reaction. There were no differences in enrichment in the quadraplex compared to performing the LM-PCR as four individual singleplex reactions, with the exception of *bla*_OXA-48-like_ (**Figure S2**).

The gene copies of ARGs prior to and after enrichment were determined by qPCR. Gene copies after enrichment were compared to the percentage of mapped reads from the same sample to determine the copies of ARGs required for adequate sequencing coverage. Based on the successful recovery of *bla*_CTX-M_, *bla*_KPC_, and *qnrS,* and the unsuccessful recovery of *bla*_OXA-48-like_, we determined that each sample needed to obtain enrichment of at least 10^10^ gc/µL to get sufficient sequencing coverage (**Figure S3**). *For bla*_OXA-48-like_, we did not obtain sufficient sequencing coverage to determine a minimum abundance for success. At this stage, primers for *bla*_OXA-48-like_ LM-PCR were redesigned and screened for improved sensitivity. The redesigned primers for *bla*_OXA-48-like_ were used in our full-scale study (**Figure S4 and S5**).

### 3.2 ARG Enrichment of ARGs and their Genomic Context from WWTPs

All four ARG targets were detected by qPCR in 100% of the wastewater influent samples (n=26) (**Figure S6)**. All samples demonstrated some level of enrichment, although the magnitude of enrichment was target dependent. *QnrS* was the most abundant enriched target (median: 1.49 x 10^12^ gc/μL; range: 7.21 x 10^10^ - 3.32 x 10^12^ gc/μL), while *bla*_OXA-48-like_ was the least enriched (median: 9.80 x 10^8^ gc/μL; range: 8.01 x 10^6^- 6.65 x 10^10^ gc/μL) across both timepoints (**Figure S7 and Table S13**). Most *bla*_CTX-M_, *bla*_KPC_, and *qnrS* targets were enriched to 10^10^ gc/µL (70-100%), which we previously determined was needed for optimal sequencing coverage. *Bla*_OXA-48-like_ demonstrated the lowest level of enrichment among treatment plants and timepoints despite utilizing the redesigned assays with enhanced amplification (<15%).

### 3.3 Unique Genomic Sequence Clusters

From sequencing LM-PCR samples, we obtained average read lengths of 1399 ± 368 bps for *bla*_CTX-M_, 1259 ± 369 bps for *bla*_KPC_, 1274 ± 387 bps for *bla*_OXA-48-like_, and 1280 ± 348 bps for *qnrS* (**Table S14**). Within each sample and ARG group, reads were aligned and clustered into highly similar sequence groups. Consensus sequences from each cluster were then generated to produce error-corrected representative sequences. After dereplication of clusters within each sample and across the full dataset (95% query coverage; 95% identity), we obtained 11 unique *bla*_CTX-M_, 7 unique *bla*_KPC_, 1 unique *bla*_OXA-48-like_, and 24 unique *qnrS* cluster variant sequences (**Figure 3 and Figure S8**). When considering the total number of clusters we obtained and the impact of sequencing effort, there was no correlation between the number of reads and number of recovered clusters for *bla*_CTX-M_ (Pearson’s r = 0.18, p-value = 0.39) and stronger correlations for *bla*_KPC_ (Pearson’s r = 0.61, p-value 0.001) and *qnrS* (Pearson’s r = 0.70; p-value <0.0001). For *bla*_KPC_ and *qnrS,* an average of four additional unique sequence clusters could be recovered for every additional order of magnitude increase in mapped reads.

**Figure 3:**
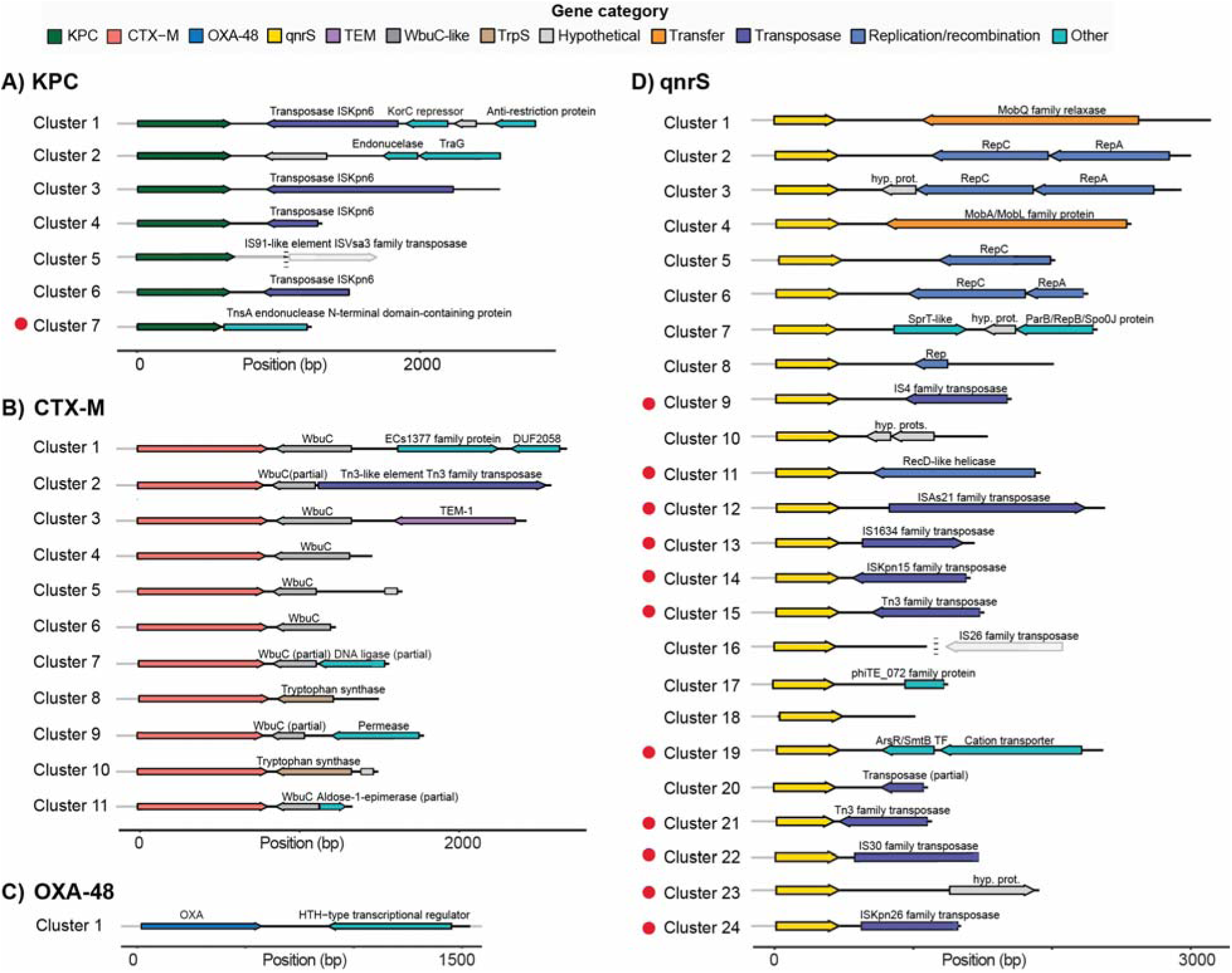
Genomic annotations of unique cluster variant sequences. Unique genomic sequence clusters captured and assembled using our LM-PCR strategy for (**A**) bla_KPC_, (**B**) bla_CTX-M_, (**C**) bla_OXA-48-like_, and (**D**) qnrS. Sequence annotations were obtained from the nearest sequence match in the NCBI database (>99% ID/coverage). Novel sequence contexts not identified in NCBI databases but recovered from multiple samples are denoted with a red circle. Annotations for novel contexts were obtained from NCBI database annotations of the cluster sequence without ARG. Position denoted is relative to the forward primer used in the amplification assay. Dashed vertical lines show downstream CDS from database hits (not part of assembly) and were included to visually distinguish similar genomic contexts.

There were 12 ‘novel’ cluster variant sequences (1 *bla*_KPC_ and 11 *qnrS*) that were not present in the NCBI nucleotide (nt) database and annotated independently from the ARG sequence. While we cannot identify the exact ARG alleles in the samples due to the partial gene coverage (based on location of primer), *bla*_KPC_ gene clusters mapped to NCBI sequences containing *bla*_KPC-2_ and *bla*_KPC-3._ *Bla_CTX_*_-M_ containing clusters in NCBI contained *bla*_CTX-M-15_, *bla*_CTX-M-55_, *bla*_CTX-M-3_, and *bla*_CTX-M-1_. *Bla*_OXA-48-like_ aligned to *bla*_OXA-48_ while the majority of *qnrS* genes aligned to either *qnrS1* or *qnrS2*.

Out of the 43 cluster variant sequences, only one cluster did not have identifiable genomic annotations beyond the ARG (*qnrS* Cluster 18). Many of the clusters identified contained genes that encode for transposases (e.g., *bla*_KPC_ Clusters 1,3,4, & 6; *qnrS* Clusters 9, 11, 12, 13, 15, 20, 22, & 24). For *bla*_KPC_ Cluster 2, a *traG* gene was identified which would be required for conjugative transfer of plasmids. Nine of 11 *bla*_CTX-M_ clusters sequences were adjacent to *WbuC*, a cupin fold metalloprotein. *Bla*_CTX-M_ Clusters 2 and 7 contain a tryptophan synthase, which encodes an enzyme that catalyzes the last two steps of L-tryptophan synthesis. *Bla*_CTX-M_ Cluster 2 also contained a Tn3-like family transposase and *bla*_TEM-1_ gene, which is another ARG that confers resistance to β-lactam antibiotics. The *bla*_OXA-48like_ cluster variant sequence mapped to an existing sequence in the NCBI database and contained an HTH-transcriptional regulator protein. Five out of 24 (20.8%) *qnrS* clusters contained a gene involved in plasmid replication. Two clusters (*qnrS* Cluster 1 and *qnrS* Cluster 4) contained a *mobQ*/*mobA*/*mobL* gene, associated with bacterial conjugation. One novel cluster contained *arsR*, related to arsenic resistance, and cation transporter genes (*qnrS* Cluster 19).

When comparing sequence clusters to each other with ARGs, there was more sequence diversity (as measured by %ID) between *qnrS* clusters (**Figure 4)** compared to *bla*_CTX-M_ and *bla*_KPC_ clusters, even when accounting for read length. *Bla*_CTX-M_ clusters were highly similar, with *bla*_CTX-M_ Clusters 1 and 3 being most distant to others. The novel *bla*_KPC_ cluster sequence, *bla*_KPC_ Cluster 7, was the least similar to other *bla*_KPC_ clusters.

**Figure 4:**
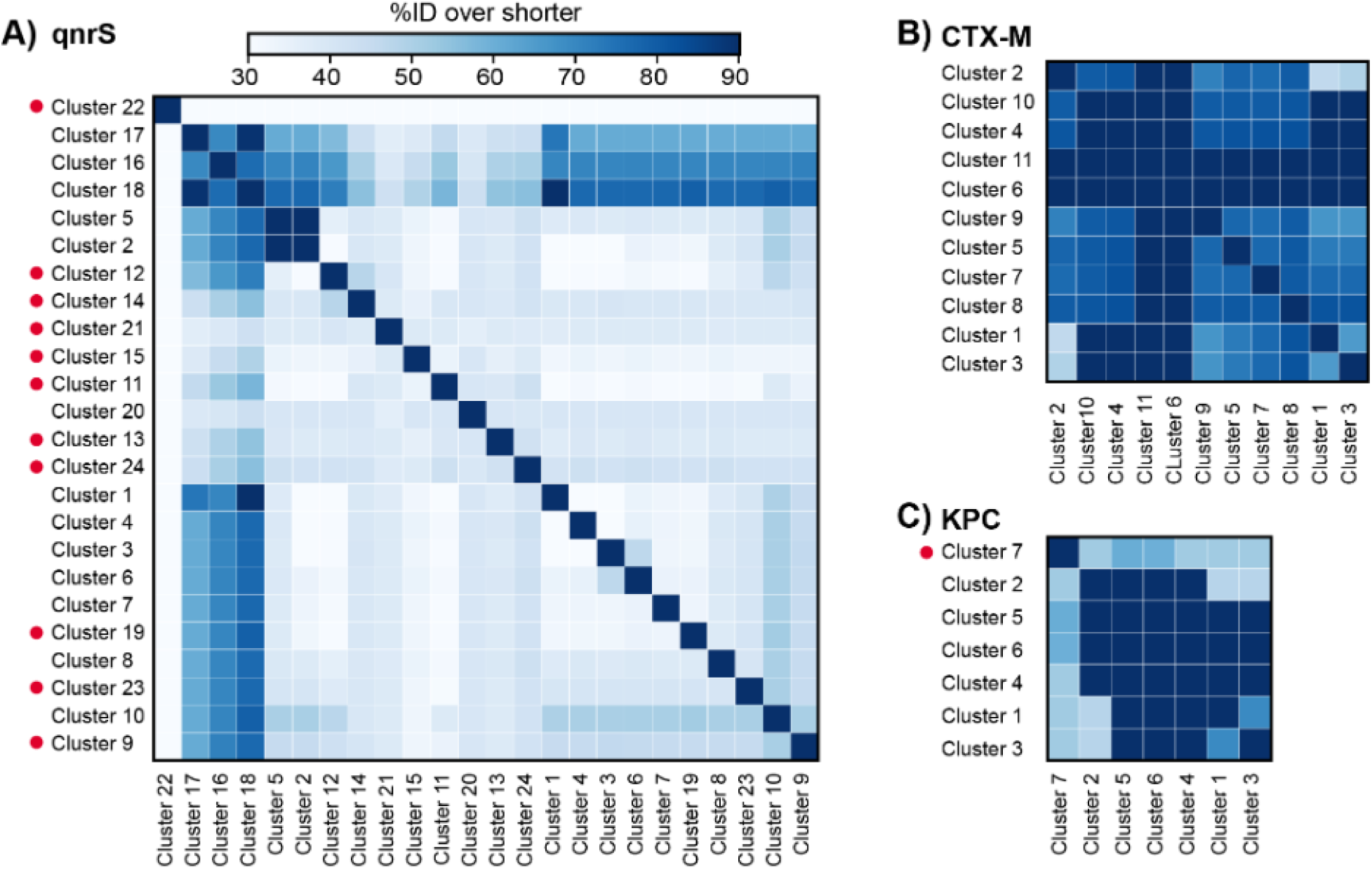
All-by-all sequence similarities of ARG-associated genomic contexts. Cross cluster sequence percent identity, for region of shared coverage (%ID over shorter), for (**A**) qnrS, (**B**) bla_CTX-M,,_ and (**C**) bla_KPC_. Clusters with novel sequence contexts not identified in NCBI databases are denoted with a red circle.

We next compared cluster variant sequences in our study to *Salmonella enterica*, *E. coli* and *Shigella* spp., and *Klebsiella pneumoniae* sequences from WA state found in NCBI’s Pathogen Detection Project database. Among *bla*_CTX-M_ and *bla*_KPC_ variants that matched Washington state pathogen genomes (**Table S15**)^44^, *bla*_CTX-M_ Cluster 2 was the most frequent, matching seven *K. pneumoniae* and seven *E. coli* isolates. In contrast, only a single *bla*_KP_ variant (*bla*_KPC_ Cluster 3) matched a WA state pathogen genome, corresponding to one *K. pneumoniae* isolate. Only one *qnrS* cluster in our study (*qnrS* Cluster 20) was found in publicly available WA State pathogen genomes. *QnrS* Cluster 20 was present in two *Salmonella enterica*, one *Klebsiella pneumoniae*, and one *E. coli* isolate.

Beyond these statewide matches, we investigated ARG cluster sequence that mapped to five or fewer records in NCBI nt database. Most of these database hits corresponded to clinical isolates sequenced through national surveillance programs spanning multiple countries (Canada, USA, China, Switzerland, Japan, and South Korea) (**Table S16).** Several *qnrS* clusters matched bacteria isolated from animal-associated sources, including domestic pig (*qnrS* Cluster 16), iridescent shark (*qnrS* Cluster 2), and largemouth bass (*qnrS* Cluster 1). A subset of clusters were reported previously in environmental samples, including river water and fecal sludge isolates (*bla*_CTX-M_ Cluster 5), hospital wastewater samples (*bla*_KPC_ Cluster 2, also mapped to clinical isolates), and wastewater effluent (*qnrS* Cluster 17). Most mapped records corresponded to plasmid sequences, consistent with the expected association of *bla*_CTX-M,_ *bla*_KPC,_ and *qnrS* with mobile genetic elements.

### 3.4 Cluster Occurrence Across Wastewater Treatment Plants

Raw reads from all WWTP samples were mapped to the dereplicated cluster variant sequences using two complementary strategies: unambiguous and ambiguous mapping. Under the unambiguous approach, each read was assigned only its single best hit, so a detected match indicates that the read is most similar to one specific cluster variant. To account for differences in sequencing effort (total reads per sample), we normalized mapped-read counts by the total number of sequenced nucleotides in that sample (**Figure 5A**). *QnrS* Clusters 1, 2, 3, and 5; *bla*_KPC_ Clusters 1, 3, and 5; and *bla*_CTX-M_ Clusters 1 and 3 show consistent detection across sample sites and timepoints, indicating that these genetic contexts are shared among geographically distinct wastewater systems.

**Figure 5:**
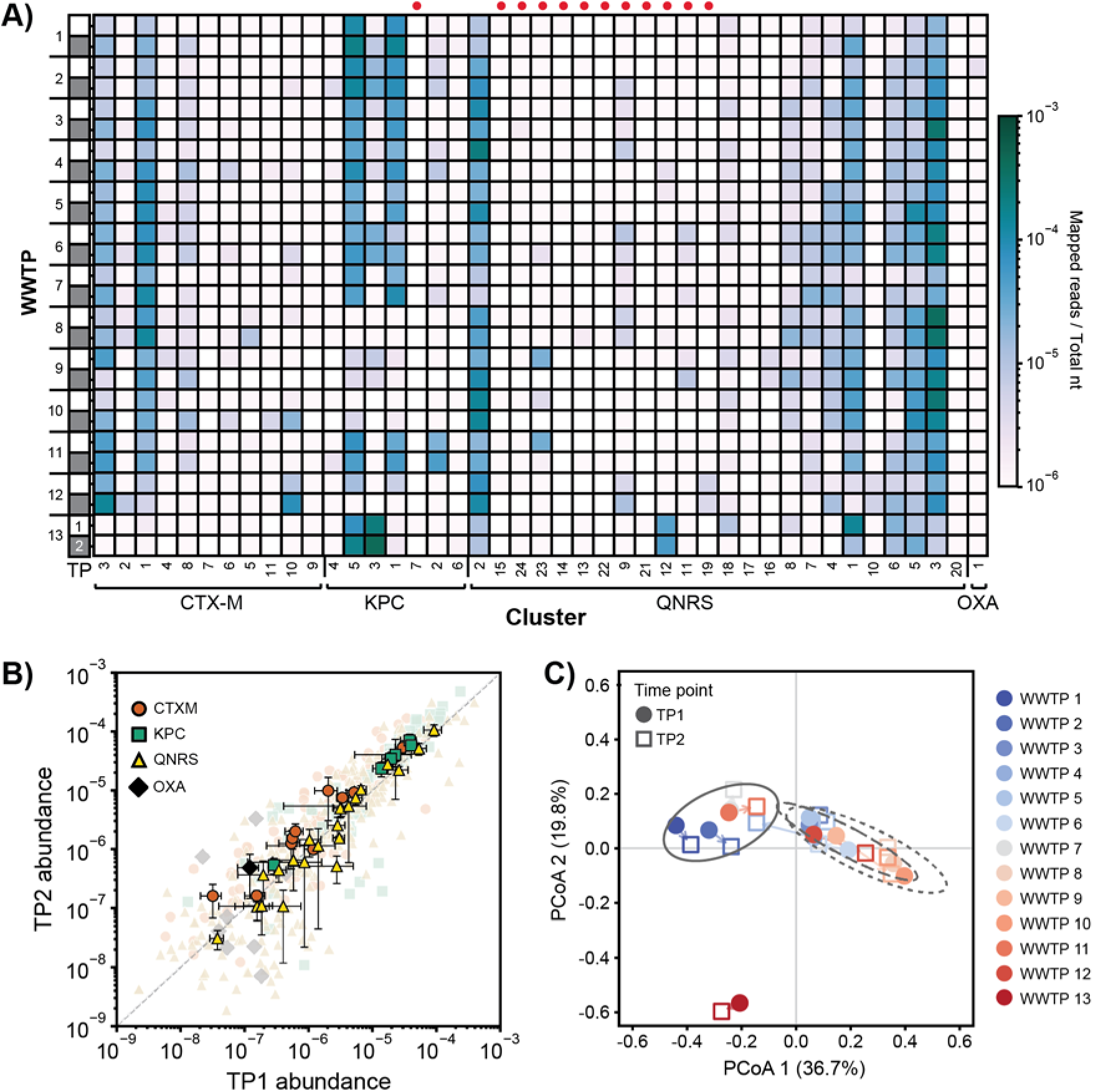
Abundance and detection of cluster variant sequences in 13 WWTP influents at two timepoints. (**A**) Raw reads for each sample were mapped to the cluster variant sequences and read coverage was normalized by total nucleotides sequenced in that sample. These reads were unambiguously mapped, meaning that a read matched to only one of the possible cluster. Novel cluster sequences, not previously found in NCBI nucleotide database, are denoted with a red dot. (**B**) The average and standard deviation (error bars) of the log-abundance of each cluster consensus sequence at timepoints 1 and 2. Each individual data point is shown in a lighter color. (**C)** Samples ordinated by Bray-Curtis dissimilarity based on normalized abundance of each ARG-containing sequence cluster. Abundances were normalized by dividing sequence read counts by total nucleotides per sample. The arrows track changes between timepoint 1 and timepoint 2 for each sample site. Ellipses show 95% confidence regions for clusters 1–3. Cluster 4 comprises only the two WWTP 13 samples, which are too few to define an ellipse.

Since read lengths varied and the longest cluster variant sequences were used as references, we repeated the same abundance analysis allowing ambiguous (secondary) mappings. Under this approach, a read may contribute to multiple cluster variants when it is compatible with more than one reference, capturing cases where short or conserved segments cannot identify a unique match. Including all mappings, *bla*_KPC_ Clusters 1, 2, and 4 and *bla*_CTX-M_ Clusters 3 and 11 were abundant in most WWTP influents (**Figure S9**). Novel cluster variant sequences tended to be present at lower abundances regardless of mapping strategy.

For any given WWTP, the log-abundances of individual ARG cluster sequences were similar in magnitude between the two timepoints (TP1 and TP2) (**Figure 5B)**. Cross timepoint analysis performed on log-abundances (n=13) shows this temporal correlation was strong for *bla*_CTX-M_ (ρ = 0.89, p-value<0.001), *qnrS (*ρ *= 0.87, p-value<0.001),* and *bla_KPC_ (*ρ *= 0.84, p-value<0.001).* Low to no correlation was observed for *bla*_OXA-48-like_ (ρ = 0.02, p-value=0.96). Higher-abundance ARG-associated clusters may disseminate more effectively in WWTPs and thus are more consistently detected over time, whereas lower-abundance clusters may exhibit greater temporal variability or fall near the detection limit of the assay. It is also possible that higher abundance clusters are associated with sewer microbiomes rather than human microbiomes.

Influent profiles separated into four groups, though with weak cohesion (partitioning around medoids; average silhouette width = 0.28; **Figure 5C**). Most WWTPs (1, 2, 5, 6, 7, 8, 9, 11,12,13) remained in the same cluster across both timepoints (*e.g.*, WWTP 1 in group 1 and WWTP 13 in group 4 at both timepoint 1 and timepoint 2). Clustering was associated with the number of hospitals contributing to the catchment areas (PERMANOVA unadjusted p-value=0.02, adjusted p-value =0.11), and WWTP flow capacity (PERMANOVA unadjusted p-value=0.04, adjusted p-value =0.12), though not when adjusting for multiple comparisons.

## 4.0 Discussion

Our targeted enrichment method recovered 43 unique ARG cluster variants representing four clinically relevant ARGs across 13 WWTPs. Many cluster variants were detected in multiple geographically distinct WWTPs, indicating that similar ARG-associated mobile genetic contexts occur across influent wastewater in Washington state. For most ARGs and WWTPs, we also observed agreement between sampling timepoints for ARG cluster variant abundances. This consistency across sites and timepoints suggests that targeted enrichment can capture reproducible ARG-context profiles in wastewater, while the extensive diversity observed, particularly among *qnrS* clusters, highlights the value of resolving ARGs within their broader genetic backgrounds.

Data regarding ARGs and their surrounding genomic context in wastewater systems remains limited compared to the extensive sequence information available from clinical isolates. Though prior wastewater metagenomic studies have characterized the genetic environments surrounding numerous ARGs using long-reads or contig assembly,^29,40,45^ these studies did not report genomic neighborhoods associated with *bla*_CTX-M_ or *bla*_KPC_. Additionally, recent targeted sequencing studies have improved recovery of *bla*_CTX-M_ from wastewater,^20,46^ but many studies focused on ARG detection rather than annotation of genomic context. In our study, one *bla*_CTX-M_ cluster variant also contained *bla*_TEM-1_, demonstrating that targeted recovery of ARG genomic neighborhoods can reveal co-occurring resistance determinants that may be missed when surveillance is limited to individual ARG detection. Prior studies of ESBL-producing isolates have suggested that co-occurrence of *bla*_CTX-M_ and *bla*_TEM-1_ may contribute to β-lactam resistance phenotypes, including through potential co-maintenance or co-expression under β-lactam exposure.^47,48^ WbuC, a putative cupin-fold metalloprotein, was identified adjacent to *bla*_CTX-M_ in nearly all *bla*_CTX-M_ cluster variants, although the downstream genomic context varied across clusters. While WbuC is not known to be directly involved in antimicrobial resistance or DNA mobilization, its recurrent association with *bla*_CTX-M_ and other ARGs has been reported previously.^49,50^

Of the four ARGs studied here, the highest diversity of genomic context was found around *qnrS* genes. Many of the genes surrounding *qnrS* were associated with DNA mobility^51^ including transposases,^23,52^ relaxases,^53^ and replicases.^54^ Other work studying *qnrS* diversity in wastewater, that used a CRISPR-Cas9 enrichment approach, also recovered previously uncharacterized *qnrS-*associated genomic contexts.^46^ Only one *qnrS* cluster in our study was found in publicly available *K. pneumoniae*, *E. coli/Shigella*, and *S. enterica* pathogen genomes from WA state. Several *qnrS* clusters had five or fewer matches in the NCBI nt database, mapping to animal-associated sources, wastewater, and clinical isolates from outside the United States. It is unknown if the *qnrS* sequences are not present in WA state pathogen genomes due to under sampling isolates or if these specific genomic arrangements have not occurred, and may never occur, in human pathogens in WA. Our results suggest that targeted wastewater sequencing can expand the observable diversity of *qnrS*-associated genomic contexts beyond those captured by isolate-based surveillance, offering a complementary view of genomic contexts circulating in environmental reservoirs.

Across all ARGs, cluster variant profiles were broadly similar within each WWTP at the two timepoints. Most cluster variants detected at TP1 (May) were also detected at TP2 (June), and cluster variant abundances were positively associated between timepoints. Previous work has documented differences in AMR composition in wastewater over shorter time periods (24-h)^55^ and season.^56^ In the approximately month-long time period of our study, PCoA analysis showed WWTPs clustered by WWTP instead of timepoints with an association, though weak, with WWTP flow rate. For future applications focused on regional or plant-level comparisons, strategically collected 24 h composite samples across seasons and at multiple WWTPs may be sufficient to capture broad differences in ARG-context profiles between populations.^57^

Together, our findings demonstrate the value of LM-PCR as a targeted strategy for resolving ARG genomic context in wastewater surveillance. Large-scale wastewater metagenomic studies have shown the importance of examining ARG flanking regions, but untargeted sequencing remains inefficient for this purpose since only a small fraction of reads (<0.05%) map back to ARGs, even when millions of reads are generated per sample.^29,40^ Other targeted approaches have improved ARG recovery, but short-read sequencing,^58^ short amplicons,^20,46,59^ or host-linkage methods such as EPIC-PCR^60,61^ are limited by sequence length or resolution of the surrounding genomic context.^62^ By contrast, LM-PCR enriched ARG-containing fragments while preserving flanking regions, allowing us to generate consensus sequences that captured both target ARGs and their associated genetic contexts. Since most of the reads obtained in this workflow contributed directly to target ARG reconstruction, LM-PCR reduced the sequencing effort spent on non-informative background reads relative to untargeted metagenomics. Combined with sample multiplexing, this approach is cost-efficient for implementation, requiring no specialized enrichment chemistry beyond commercially available oligos and enzymes for long-range PCR. These features make LM-PCR a promising, practical, and scalable method for integrating ARG-context resolution into our wastewater AMR surveillance programs.

Despite these advantages, broader implementation of LM-PCR for wastewater ARG surveillance will require attention to several technical and methodological constraints. First, although we included bead-based size-selection steps, both PCR and nanopore sequencing can be biased toward shorter DNA fragments, which limits the upper range of recoverable sequence context. Future work could explore additional size-selection strategies, such as gel extraction or BluePippin-based selection, to enrich for longer fragments. Additionally, during method development, we found that it was necessary to include experimental steps to minimize the formation of ARG concatemers, which can arise during ONT’s SQK-LSK114 library preparation, and bioinformatic steps to split concatemeric reads. When multiplexing samples, omitting these steps could interfere with cluster formation and increase the risk of false-positive sample assignments or genetic context assignments. Another limitation of the approach is that the recovered sequence context is directional. In this work, we only captured sequence context 3’ of the ARG coding sequence. A second round of targeted sequencing using the same general strategy, but with primers targeting the antisense strand, could be used to recover 5’ flanking regions. However, linking the corresponding 3’ and 5’ regions would remain challenging, particularly when highly similar ARG sequences are present since they can create ambiguity in assigning flanking regions to the same ARG. Finally, we note that this wastewater-based detection approach cannot distinguish among potential sources, including human microbiome shedding, animal microbiome shedding, or bacterial proliferation within the sewer system.^63^

## Contributions

Project conceptualization was performed by ERF, AJP, RSK, and BM. Methodology for this work was developed by MO, ERF, SK, CM, DG, AJP, and RSK. Sample coordination was performed by PB, EK, MF, and BM. Data analysis was performed by MO, ERF, and JAM. Additional data was provided by BA and ED. Subject matter expertise was provided by KK. Visualization of data and results was performed by MO, ERF, and JAM. Funding for this work was acquired by ERF, AJP, RSK, and BM. This project was administered and supervised by ERF. Writing of original draft was carried out by MO, ERF, and JAM. Reviewing and editing of the manuscript was performed by all.

## Competing interests

The authors of this work declare no other competing financial interests.

## Data availability

Sequencing data is available in the SRA under BioProject PRJNA1514801.

## Code availability

Code used for data processing and analysis is available in the public fuhr-microlab/lmpcr_arg repository on GitHub.

## Acknowledgements

This work was supported by U.S. Centers for Disease Control BAA 75D301-22-R-72097 and in part by the Epidemiology and Laboratory Capacity for Infectious Diseases Cooperative Agreement (grant number 24NU51CK000364). JAM was supported by the National Science Foundation Grant MCB-2440857. AJP is a Biohub, San Francisco Investigator. The funders had no role in study design, data collection and analysis, decision to publish, or preparation of the manuscript. The authors gratefully acknowledge the operators and staff at the participating wastewater treatment facilities for their invaluable support in collecting samples. We would also like to thank the Department of Community Health-Epidemiology, Walla Walla County; Lianne Bradshaw, Epidemiologist at Kittitas County Public Health Department, Ellensburg, WA; Neil Panlasigui, Epidemiologist, Skagit County Public Health; Danielle Lee, Communicable Disease Nurse Clinical Supervisor, Skagit County Public Health; Alex Gee, Senior Manager, Skagit County Public Health; Public Health – Seattle & King County; Snohomish County Health Department; Josina Bickel, Disease Control Investigator at Yakima Health District; Jefferson County Public Health; Disease Prevention & Response, Spokane Regional Health District; Communicable Disease Division, Benton-Franklin Health District; and Public Health-Island County.

## Notes

### Competing Interest Statement

The authors have declared no competing interest.

